# Wastewater Analysis of SARS-CoV-2 as a Predictive Metric of Positivity Rate for a Major Metropolis

**DOI:** 10.1101/2020.11.04.20226191

**Authors:** L.B. Stadler, K.B. Ensor, J.R. Clark, P. Kalvapalle, Z. W. LaTurner, L. Mojica, A. Terwilliger, Y. Zhuo, P. Ali, V. Avadhanula, R. Bertolusso, T. Crosby, H. Hernandez, M. Hollstein, K. Weesner, D.M. Zong, D. Persse, P.A. Piedra, A.W. Maresso, L. Hopkins

## Abstract

Wastewater monitoring for SARS-CoV-2 has been suggested as an epidemiological indicator of community infection dynamics and disease prevalence. We report wastewater viral RNA levels of SARS-CoV-2 in a major metropolis serving over 3.6 million people geographically spread over 39 distinct sampling sites. Viral RNA levels were followed weekly for 22 weeks, both before, during, and after a major surge in cases, and simultaneously by two independent laboratories. We found SARS-CoV-2 RNA wastewater levels were a strong predictive indicator of trends in the nasal positivity rate two-weeks in advance. Furthermore, wastewater viral RNA loads demonstrated robust tracking of positivity rate for populations served by individual treatment plants, findings which were used in real-time to make public health interventions, including deployment of testing and education strike teams.

## Introduction

Wastewater monitoring for severe acute respiratory syndrome coronavirus 2 (SARS-CoV-2), the causative virus of novel pneumonia (COVID-19), represents a paradigm shift for real-time monitoring of community infection dynamics (*1, 2*). Although primarily considered a respiratory disease, surveillance of SARS-CoV-2 in wastewater is possible because infected individuals also excrete SARS-CoV-2 in their stool. SARS-CoV-2 RNA has been detected in the stool of ∼ 40% of positive individuals (*3*). Diarrhea is a common symptom in those infected, and can often be a leading symptom and/or the only symptom present (*4*). Finally, SARS-CoV-2 efficiently replicates in an intestinal tissue model (*5*), raising the possibility that the intestines may also become infected by SARS-CoV-2, thereby implicating wastewater as an effective tool for disease surveillance.

SARS-CoV-2 RNA has been detected in numerous wastewater samples across the world (*6*– *15*). Wastewater monitoring for SARS-CoV-2 has numerous benefits as a complementary tool to diagnostic testing: it is a cost-effective means to surveil a significant portion of the US population served by public sewer systems; it could identify outbreaks earlier than diagnostic testing (*6, 7, 16*); it does not require individuals to opt-in to participate and thus may capture the substantial undocumented infections and spread of the virus (*17*); it captures both symptomatic and asymptomatic infections of SARS-CoV-2 (*18, 19*); it could be applied in resource-poor communities with limited access to healthcare facilities; and it may be used to determine novel associations between viral transmission, clinical burden, and population demographics. While there is interest in implementing wastewater monitoring on a national scale as a lead-indicator for SARS-CoV-2 infections and disease prevalence, the relationship between viral signal and key community metrics such as the positivity rate (which has been used to make policy decisions) is poorly characterized. Furthermore, the impact of assessing viral RNA levels with enhanced spatial resolution (i.e., unique geographic sites within a large area) as an indicator of disease prevalence or outbreaks is not known.

Here, we conducted an extensive wastewater monitoring program for SARS-CoV-2 for a city of 3.6M people (Houston, Texas) that underwent a massive surge in cases, hospitalization, and deaths over a 22-week period from May to October of 2020. By collecting samples from 39 wastewater treatment plants (WWTPs) across Houston that treat a combined average flow of 250 million gallons of wastewater per day, we demonstrate with nonparametric regression models that changes in SARS-CoV-2 RNA levels in wastewater provide a two-week lead-indicator for changes in positivity rate. The 39 WWTPs sampled range in size from 0.11 to 80 million gallons per day (MGD), corresponding to service population sizes of approximately 5,100 to 564,000, thus providing an array of population sizes by which to compare datasets and results. In addition, this geographic resolution in sampling was used to identify regional hotspots which allowed public health officials to mobilize strike teams to increase testing, education, and contact tracing in soon-to-be affected areas.

## Results and Discussion

### SARS-CoV-2 viral RNA loads correlate with clinical positivity rates

Between May 11 and October 10, 2020 we collected and analyzed 24-hour time-weighted composite influent samples from between 16 and 39 wastewater treatment plants (WWTPs) (Table S1) once per week that collectively serve over 3.6M people in Houston (**Fig. 1A**). Samples were collected from all sites on the same day each week (Tuesday mornings, corresponding to a 24 hour composite that ran from 7am Monday to 7am Tuesday). To increase the rigor and reproducibility of the analysis, SARS-CoV-2 RNA was quantified from influent wastewater samples in two independent laboratories: Baylor College of Medicine and Rice University. Viral loads for each WWTP were calculated by multiplying the measured virus RNA concentration by the 24-hour average flow rate for the corresponding WWTP. We applied a multi-level flexible model that took into account quantification of SARS-CoV-2 RNA using two primer sets (N1 and N2, the same primer sets used by the CDC 2019-nCoV RT-PCR diagnostic test), results reported by two different laboratories, and concentration replicates analyzed at each laboratory. Nonparametric regressions were fit to weekly average viral load data for each individual site and to the aggregate viral load (**Fig. 1B**). **Figure 1B** includes only the 16 WWTPs that were sampled between May 11 and October 5. Additional plots including all sites are provided in **Fig. S2** and **S3**.

**Fig. 1.**
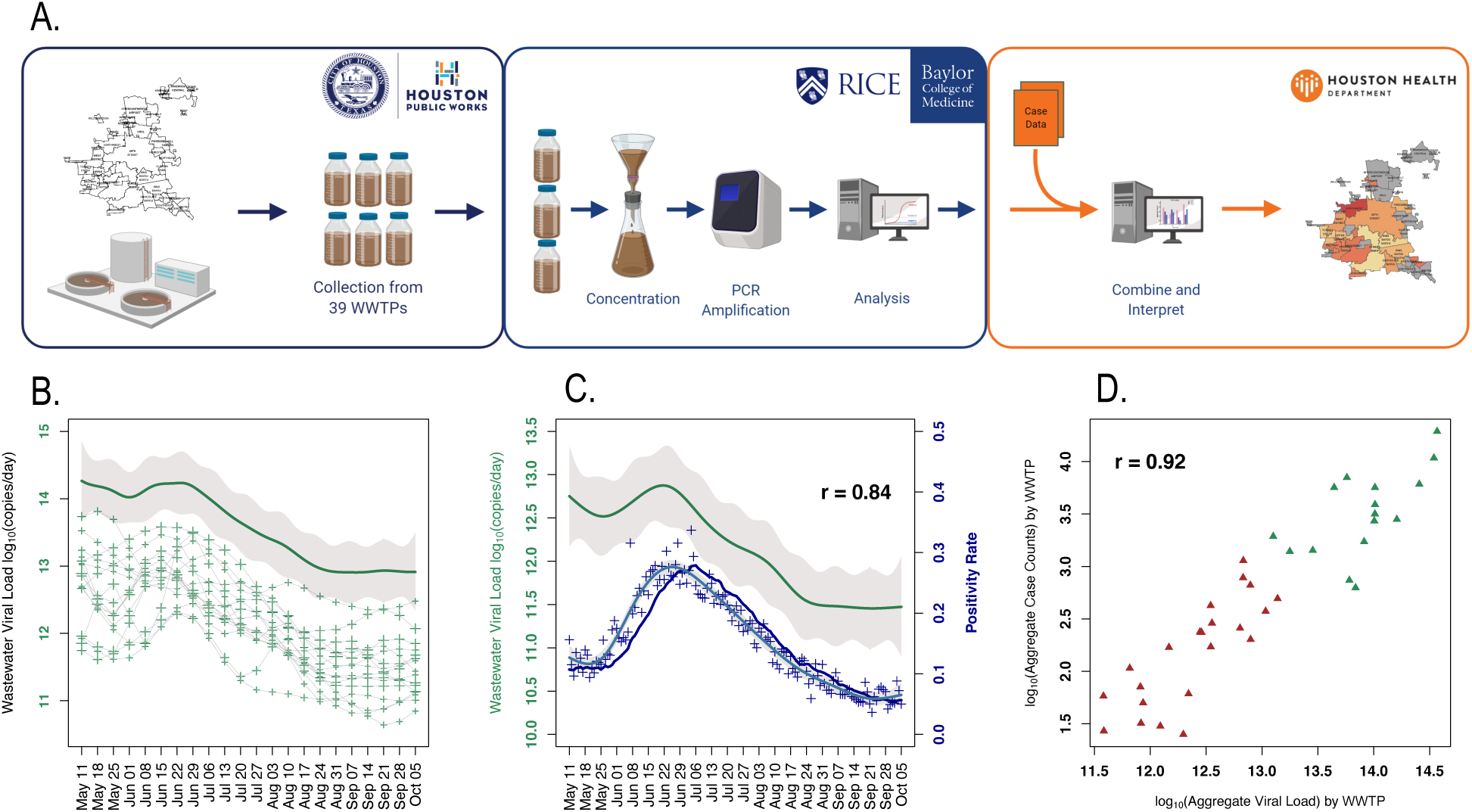
Wastewater monitoring in Houston shows that SARS-CoV-2 levels track positivity rate. (**A**) Overview of weekly SARS-CoV-2 wastewater surveillance system. (**B**) A nonlinear regression (spline) was fit to the observations from each WWTP (+ symbols connected by lines represent the same WWTP; the size of the + denotes the level of uncertainty). The individual splines were inverse log10 transformed, summed, and then 1og10 transformed to form the overall spline (green line). Grey line is the 95% confidence band for the overall estimate derived from the sum of the variances (each spline). (**C**) Green line is the averaged spline for the wastewater viral levels for the 16 WWTPs (from panel B). Dark blue line is the 14-day moving average of the positivity rate (+ denoting the daily observations; the light blue line shows the nonlinear regression (spline) fit to those observations). Grey represents the 95% confidence bands. (**D**) Log10 total positive clinical cases against log10 total viral load over the study period. Symbols denote individual WWTP’s positive cases and total viral load. Green symbols denote wastewater viral loads and cases (May 11 - October 5) and red symbols sites between July 8 and October 5.

Houston underwent a major surge in cases, hospitalizations, and deaths that began in the first week of June, peaked in late July, and has trended downward since then (**Fig. 1C** shows the positivity rate as an example). At one point, the city averaged more than 1,500 daily cases, representing one of highest rates of daily case growth in a metropolitan area in the U.S. In the beginning stages of sampling prior to the clinical surge, more than half the sites sampled (16 total at this point) were positive for SARS-CoV-2. Although unaware of the fact at the time, between May 11 and 18, we recorded the second highest levels of viral load behind only the peak viral load detected during the surge at the end of June/early July. Following May 25, a stark drop in the mean viral load was observed across all sites. However, in early June, the case number began to increase rapidly, which, incidentally, was preceded by a concomitant rise in viral RNA levels in the wastewater, that peaked in late June.

The wastewater viral load tracked the positivity rate (based on specimen date) for the 16 WWTPs that have been analyzed since May 11 and serve a total population of 2.7M (**Fig. 1C;** Spearman r = 0.84, the correlation was performed at the weekly level between the wastewater and positivity rate splines). Wastewater viral load and positivity rate data were smoothed using nonparametric regressions to significantly improve the strength of the correlation between the datasets. SARS-CoV-2 RNA wastewater loads are subject to variability for numerous reasons, including the characteristics of the infected individuals contributing virus to wastewater in their stool, variability in wastewater flows, autosampler aliquots that comprise the composite sample, and variability in processing methods used to concentrate, extract, and quantify the viral RNA. Similarly, positivity rate data are susceptible to variability in the number of individuals tested each day. Smoothing effectively de-noised the two imperfect and highly variable datasets.

A cross-correlation analysis was used to assess whether a time displacement of one dataset relative to the other impacts the strength of the correlation between the two. We observed a strong cross-correlation up to fourteen days (Spearman r > 0.8 for smoothed datasets and r > 0.6 for raw datasets), suggesting that detection of SARS-CoV-2 RNA in wastewater could serve as a leading indicator of the positivity rate for community recorded infections. Previous studies have reported the detection of SARS-CoV-2 RNA in influent wastewater prior to known clinical cases in sewersheds (*6, 7, 20, 21*). Peccia et al. measured SARS-CoV-2 RNA in primary settled solids and found that concentrations of SARS-CoV-2 RNA in the solids were 0-2d ahead of SARS-CoV-2 positive test results (by date of specimen collection) (*16*). Many WWTPs do not have primary treatment, including all of the WWTPs in Houston, TX. In addition, for cities with large centralized systems, community-level monitoring will require sample collection from sewer lines. Thus, these approaches will require measuring virus from untreated wastewater, where viral concentrations are typically present at lower concentrations than in primary solids (*22*). Our findings show significant associations between influent wastewater concentrations and positivity rates and suggest that influent SARS-CoV-2 RNA concentrations may lead positivity rates by 7 to 14 days in such samples.

Total viral load for each WWTP was plotted against the total clinical positive cases for the corresponding service area for the entire study period (**Fig. 1D**). Combining all 39 sites sampled over the entire 22-week study period revealed a strong relationship between the wastewater viral load and positive cases (Spearman r = 0.92). This provides compelling evidence that wastewater monitoring of SARS-CoV-2 RNA could be used to estimate COVID-19 disease prevalence in communities, in addition to a tool for trend tracking. There is considerable uncertainty in converting instantaneous wastewater viral loads of SARS-CoV-2 RNA to number of infected people due to variability in RNA excretion rates between individuals and over time. Our results indicate that this conversion may be possible as has been shown with polio (*23*), as better estimates of population-level fecal shedding rates of SARS-CoV-2 are established, and losses of SARS-CoV-2 RNA during sewer transport and during sample processing are quantified.

### Correlation between wastewater viral loads and positivity rates was robust for individual WWTPs

We assessed the correlation between wastewater viral RNA levels and positivity rates for each individual WWTP sampled. For the 16 WWTPs of which sampling began on May 11, we observed significant correlations between wastewater viral loads and positivity rates (Spearman r = 0.49 – 0.96 for smoothed datasets, 0.45 – 0.89 for raw datasets; **Fig. 2**). There was no relationship between the strength of the correlation between the wastewater viral load and positivity rate regressions and WWTP flow rates, service populations, or geographic areas served. For the additional WWTPs that have been sampled since July 8, 2020, we largely observed that wastewater viral loads tracked positivity rates (Spearman r = 0.52 – 1.0 for smoothed datasets, 0.08 – 0.86 for raw datasets; **Fig S3**), where there was sufficient clinical testing data available to compare to the wastewater data. In many of the sewersheds, there was insufficient clinical testing performed to estimate a positivity rate (here, positivity rates were only calculated if at least four diagnostic tests per week were performed in the sewershed service population). Thus, the lack of correlation may be due more to the lack of robust nasal testing rather than signal of virus from the wastewater. The largest WWTP sampled, the 69^th^ Street WWTP, is representative of a typical centralized WWTP that serves a large, urban area, whereas the smallest WWTP sampled is similar in size to the thousands of WWTPs that serve smaller, rural and suburban communities. Of note, strong correlations were continuously observed as wastewater viral levels and the positivity rate continued to decline.

**Fig. 2.**
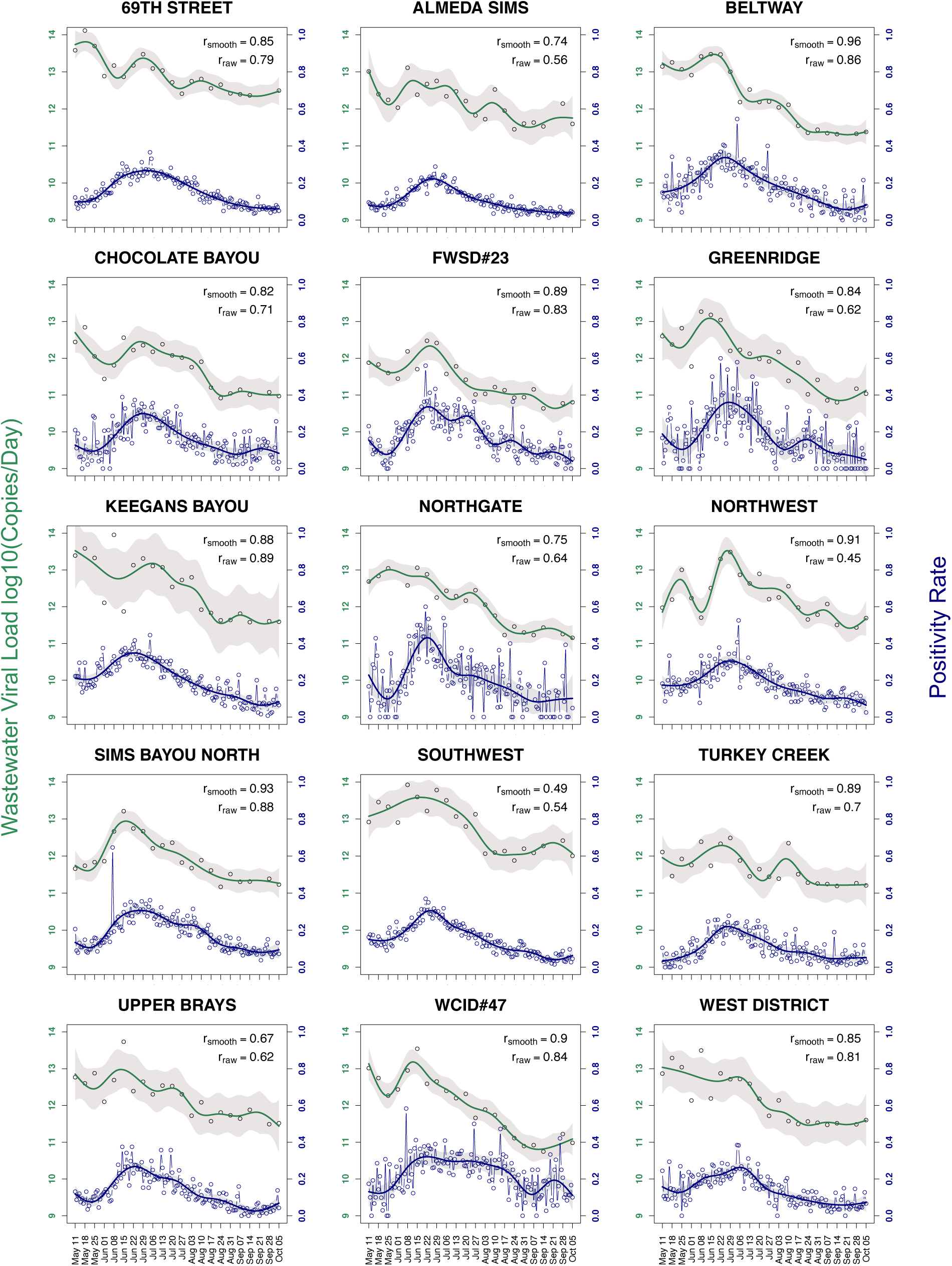
Wastewater viral loads and positivity rates for 15 individual WWTPs sampled between May 11 and October 5. Individual observations for the wastewater viral load and positivity rate are denoted by _°_ and green (wastewater) and blue (positivity rate) lines are the nonlinear regressions (splines) fit to the observations. Grey represents the 95% confidence bands. r_smooth_ is the Spearman correlation estimate (r) between the wastewater and positivity rate splines taken weekly, and r_raw_ is between the raw observations taken weekly on the dates with wastewater observations. A total of 16 WWTPs were sampled, but two WWTPs, Sims Bayou North and Sims Bayou South, have overlapping geographic service areas, so the wastewater data for both WWTPs was compared to clinical positivity rate data for the combined population served by the facilities and only Sims Bayou North is shown.

### Predictive models forecast positivity rate from wastewater viral load

The strong leading relationship of wastewater viral loads and clinical positivity rate provides the basis for a predictive model of positivity rate from observed wastewater viral load. We considered two models to predict the smoothed daily positivity rate. The first model uses the current, one- and two-week prior data of the average observed wastewater viral load (in copies day^-1^). The second model uses only one- and two- week prior data to provide a true one-week predictive model. Wastewater viral loads were log10 transformed before fitting. Each model accounts for the WWTP and the temporal structure of the data. The in-sample Spearman correlation between observed and predicted positivity rate is 0.83 for model 1 and 0.79 for model 2 (**Fig. 3A**). Using the nonparametric estimate of wastewater viral load as input, the predictive models were used to generate estimated positivity rates and evaluated against the smoothed positivity rate (**Fig. 3B** and **Fig. S4** show the time series plots for the model estimates overlaid on the positivity rate for individual WWTPs).

**Fig. 3.**
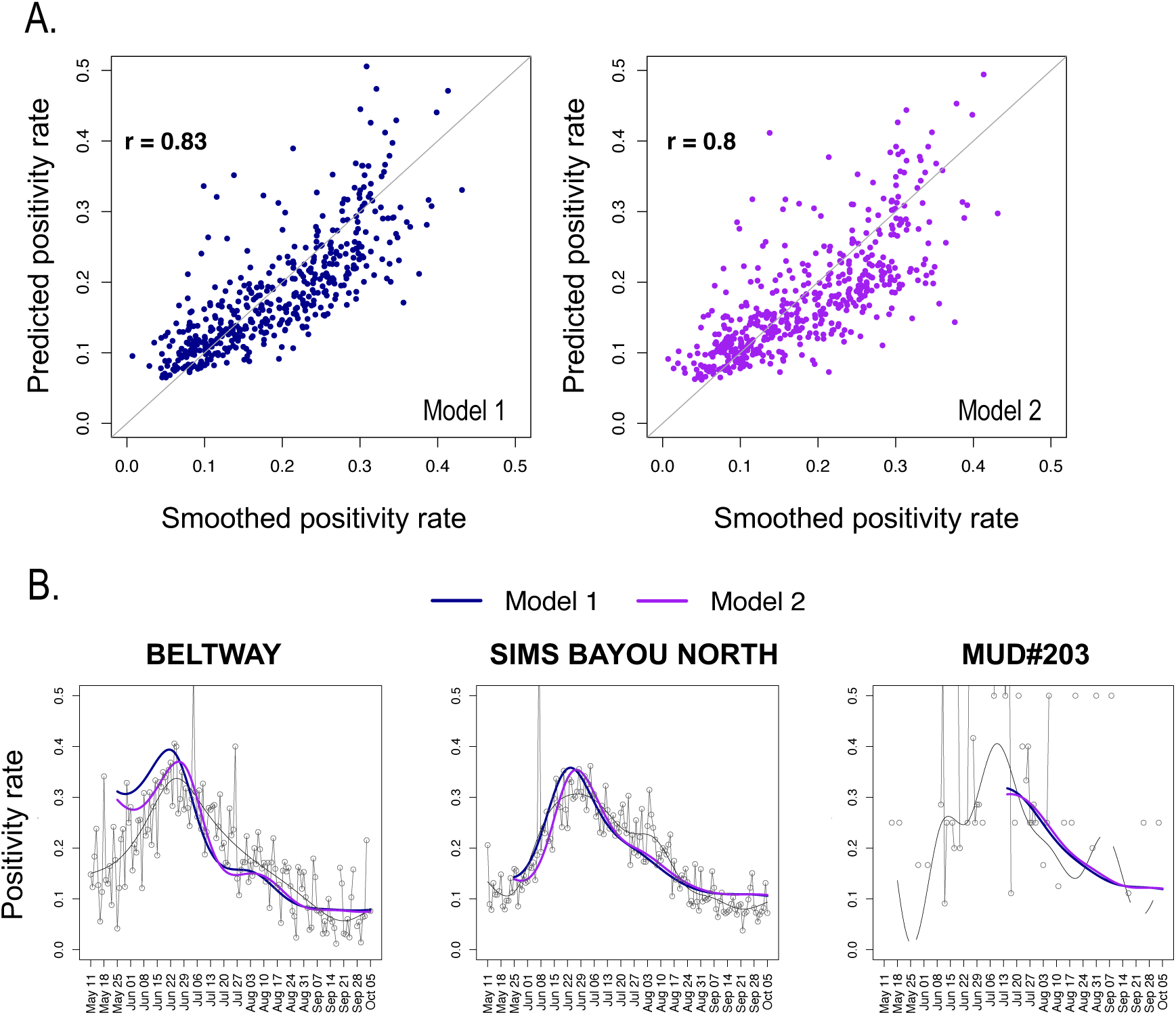
Wastewater viral loads predict positivity rates. (**A**) Predictive Model 1 uses information about the WWTP and wastewater viral load data from the current week and two previous weeks to estimate the smoothed positivity rate for the WWTP service area. Model 2 is a true one-week predictive model and uses information about the WWTP and the two previous weeks of wastewater viral load data to predict the positivity rate for the service area. The plots show the predicted positivity rate versus the smoothed clinical positivity rate (Model 1: Spearman r = 0.83; Model 2 Spearman r = 0.79). Each dot represents a weekly positivity rate from an individual WWTP. WWTP positivity rate is considered missing, and excluded from the analysis, if 4 or fewer tests results are provided in a day. (**B**) Comparison of Model 1 (blue) and 2 (purple) predicted positivity rates and clinical positivity rates for three individual sewersheds over the study period. Daily clinical positivity rates are shown as grey circles and the smoothed positivity rate is represented by the grey line.

Given the strong predictive relationship of wastewater viral loads and positivity rates, wastewater monitoring represents a viable approach to test the entire population of Houston simultaneously and a means to provide continuous, weekly monitoring of the entire population. This is particularly critical if diagnostic testing rates decline. In 12 of the 39 sewersheds monitored, there was at least one occurrence of when less than 0.1% of the sewershed’s population was nasal tested over a 7-day period during our study period. These 12 sewersheds serve a total population of 613,000, approximately 15% of the total population of Houston. In these sewersheds, wastewater monitoring represented the primary means of assessing the magnitude of disease impact in the community. For example, when applied to the sewershed MUD#203 (**Fig. 3B**) the predictive model could be used to estimate positivity rate during periods when clinical testing data was sparse. This approach illustrates the power of using wastewater viral load to forecast positivity rates in communities. While the models shown here are specific to the Houston system, this model structure could be applied to other cities to forecast positivity rates based on measured viral loads.

### Rate of change of viral levels identifies outbreaks for public health action

Time-series geospatial analysis of population normalized wastewater viral loads can be used to identify locations experiencing high infection burden each week (**Fig. 4A** and **Fig. S5**). To identify specific areas that are experiencing rapid community spread of the virus, we can visualize the change in the wastewater viral load from week to week. Assessment of the rate of change allows one to determine how an outbreak is accelerating with time, a critical metric when considering the efficient application or exhaustion of intervention or healthcare resources. When the time-series for the entire survey period is transformed into a heat map for all 39 sites, clear week to week acceleration and deceleration of viral levels are observed (**Fig. 4B**). Remarkably, every site across the entire city, albeit 1, showed a net increase in viral levels during the June 29 to July 6 week interval (**Fig. 4B**, red band on heat map). It is noteworthy that the total number of hospitalizations in Houston peaked the week of July 14 – 21, indicating that the significant acceleration of viral levels during early July as indicated by wastewater analysis likely drove the major clinical surge Houston experienced later in the month. Furthermore, the rate of change in viral levels provides critical information for healthcare officials because it pinpoints areas where infections are most rapidly worsening or improving. For example, between September 7^th^ and 14^th^, while the viral load across the city appeared to plateau (**Fig. 1B**), there were areas that experienced significant increases and areas with significant decreases in viral load (**Fig. 4B**). In this specific week, the areas with significant increases in viral loads, indicative of community spread, were distributed across the city. This reflects the reality that disease burden is heavily influenced by local outbreaks that can be heterogeneously distributed across the city.

**Fig. 4.**
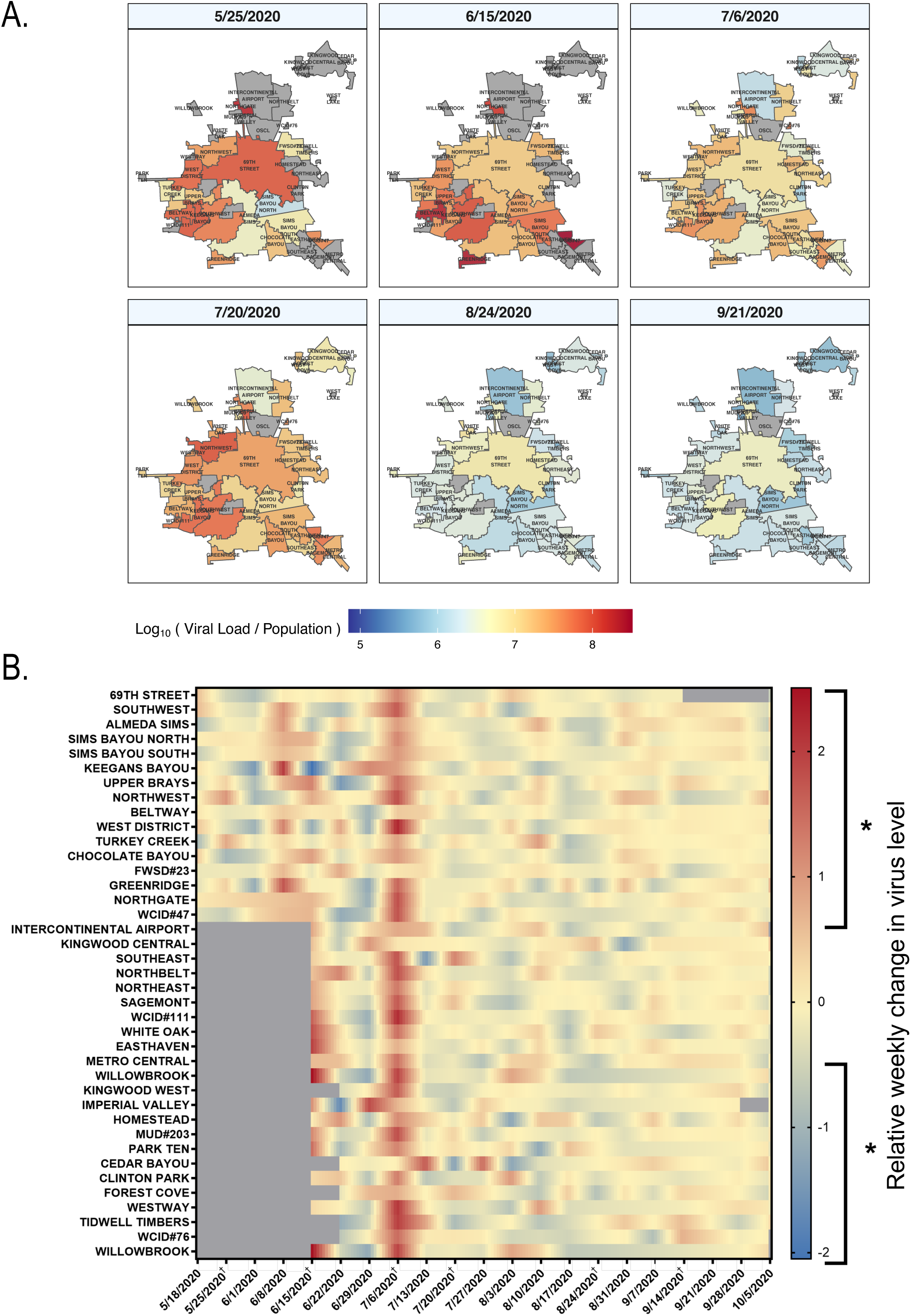
Identification of geographic areas of concern based on population normalized SARS-CoV-2 wastewater load (log10(copies/person)) and weekly change in viral load. (**A**) WWTP service areas are outlined in black. Weekly viral loads were normalized by service population. Panels depict six time-points across the 22-week study period corresponding to before, during, and after the surge in positive cases in Houston. Grey areas indicate service areas that were not sampled or with missing data that week. (**B**) Heatmap of the direction, magnitude and significance of the one week change in log10(virus copies/day). A regression model was fit to measure the change from week (n-1) to week (n). Red colors indicate an increase in viral load since the previous week, while blue indicates a decrease. Grey areas indicate service areas that were not sampled or with missing data that week. The 16 WWTPs where sampling started in May are at the top of the heatmap ordered by size of population serviced. Remaining WWTPs follow, again ordered by size of population serviced. Brackets labeled with * on the scale represent values where at least 95% of the trend coefficients had a p-value < 0.05. Dates shown in panel A are denoted with †.

The wastewater data was used to inform Health Department interventions. The WWTP that serves each city zip code, or the majority of the zip code, was identified. The WWTP virus load and the one week trend data for the corresponding zip codes, along with zip code level clinical positivity rate, were evaluated by a committee of Houston Health Department public health professionals, including the directors of the Office of Planning Evaluation and Research for Effectiveness, Office of Chronic Disease, Health Education and Wellness, and the Public Health Preparedness, Disease Prevention and Control Division. The committee uses the data to identify the15 highest priority zip codes for public health intervention and then deploys strike teams to provide education and increased free clinical testing in these zip codes. Between August 15 and September 15, 2020 targeted intervention in these zip codes resulted in educating 74,000 individuals. In light of a recent decreasing trend in clinical test penetration, wastewater data are becoming increasingly more important for virus surveillance.

Wastewater monitoring represents a rapid, inexpensive approach to obtain comprehensive coverage of large populations. Here, longitudinal sampling over 147 days of 39 wastewater treatment plants that vary in flow, population served, and geographic service area led to the discovery of significant correlations between SARS-CoV-2 RNA levels in wastewater and positivity rates across numerous zip code areas captured by the WWTPs. Based on the strength of the correlation, we show that wastewater viral loads can be used to predict positivity rates two-weeks in advance. Furthermore, granular detection of SARS-CoV-2 in sewersheds can be used to identify local outbreaks which here allowed prioritization in real-time of the allocation of healthcare resources, including free diagnostic testing and education. Additional research is needed to estimate disease prevalence from wastewater. This includes knowledge of SARS-CoV-2 RNA titers in stool of symptomatic and asymptomatic people, a quantitative characterization of losses of viral RNA during sewer transport due to dilution, adsorption, and degradation and the development of systematic approaches for selecting monitoring locations based on the above parameters. We propose the approaches developed here can be used to develop a regional, national, and/or global wastewater surveillance program for SARS-CoV-2 with similar success as observed for Houston and that might also translate to other respiratory or gastrointestinal viruses. After a vaccine is developed and administered, wastewater monitoring can facilitate early detection of re-emergence of SARS-CoV-2 in communities and be used to direct resources for vaccine administration, in line with previous work performed in the 1960s that facilitated the accelerated delivery of the oral polio vaccine to prevent outbreaks.

## Materials and Methods

### Sample collection and wastewater treatment plant (WWTP) flow data

Samples were collected from 39 different WWTPs within Houston. The WWTPs cover a total service area of approximately 580 square miles. Information for each WWTP including average flow rate, sewershed area, and service population are provided in **Table S1**. Employees of Houston Water first collected samples from refrigerated, 24-hour time-weighted, composite samplers that drew directly from the influent channels at each WWTP. Average influent flow rates over the 24-hour collection period were provided by Houston Water for each facility sampled.

After collection from each WWTP site, samples were transported to Houston Water’s central laboratory facility on ice, aliquoted into bottles, immediately placed back on ice, and then transported to laboratories at Baylor College of Medicine (BCM) and Rice University (Rice) for analysis. In both laboratories, samples were stored at 4 °C prior to analysis for no longer than 36 hours. SARS-CoV-2 RNA quantification methods were developed based on previous studies (*24*–*27*). Methods differed between BCM and Rice laboratories, and evolved within our individual laboratories over the course of the study period, as detailed below.

### Virus concentration

#### BCM concentration methods

Between May 11 and June 29, 2020, SARS-CoV-2 in wastewater samples was concentrated using a PEG precipitation method in the BCM laboratory. Wastewater samples were aliquoted into 200 mL triplicates then centrifuged at 7,140 g for 15 minutes at 4 °C in 500 mL polypropylene bottles to remove sludge and large debris. Supernatants were passed through 0.22 µm filters (SCGPS05RE, MilliporeSigma), transferred to clean polypropylene bottles, and precipitated with PEG (8% w/v, 16 g) and NaCl (0.5 M, 5.844 g) overnight at 4 °C. Precipitated filtrates were then centrifuged at 16,900 g for 30 minutes at 4 °C, supernatants poured off, and pellets resuspended in 2 mL 1X PBS solution.

On July 6, Baylor College of Medicine (BCM) switched to concentrating SARS-CoV-2 in wastewater samples using an electronegative filtration method. We aliquoted wastewater samples into 40 mL triplicates then gently centrifuged at 3,000 g for 1 minute in 50 mL conical tubes to remove sludge and large debris. A 6-head (EZFITMVHE3, MilliporeSigma) EZ Fit Manifold (EZFITBASE6, MilliporeSigma) was employed to pull 25 mM MgCl_2_ supplemented, 25 mL samples through 0.45 µm pore size, electronegative microbiological analysis HA filters contained within EZ Fit Filtration Units (EFHAW100B, MilliporeSigma) to bind RNA/virus. The funnel was then disassembled and the filter flipped over. The funnel was reassembled with the inverted filter, and positive pressure applied to elute virus components off the filter with 5 mL of 1 mM NaOH. 2.5 mL of eluent was collected in a 15 mL conical tube and neutralized with 12 µL of 100 mM H_2_SO_4_.

#### Rice electronegative filtration concentration

Between June 22 and August 17, 2020, SARS-CoV-2 in wastewater samples was concentrated using an electronegative filtration method. Wastewater (50 mL) was aliquoted into a 6-head, Multi-Vac 610-MS Manifold (180310-01, Sterlitech) containing a pre-DI-washed 0.45 µM pore size, electronegative microbiological analysis HA filter (HAWG047S6, MilliporeSigma). On August 24th, we switched to centrifuging the influent samples to remove solids prior to filtration. This change was implemented because it reduced filtration time and did not significantly impact the measured concentrations of N1 and N2 in samples (data not shown). Influent wastewater samples were centrifuged for 10 minutes at 4,100 g and 4 °C. Subsequently, 50 mL of supernatant was poured into the vacuum manifold followed by the addition of MgCl_2_*6H_2_O to achieve a final concentration of 25 mM. The samples were gently swirled with a pipette tip to homogenize and allowed to sit for five minutes. A vacuum pump then pulled the sample through the filter. After filtration was complete, the filter was folded and placed into a bead tube containing 0.1 mm glass beads. Bead tubes containing filters were stored at −80 °C and allowed to freeze prior to bead beating and nucleic acid extraction.

### Nucleic acid extraction

#### BCM nucleic acid extraction

Between May 5 and June 12, 2020, viral RNA from wastewater eluates and precipitates was extracted using the QIAamp Viral Mini RNA Kit (52906, Qiagen) with the QIAcube Connect (9002864, Qiagen) automated platform, according to manufacturer instructions. 140 µl of wastewater was extracted and eluted with 100 µl of elution buffer (May 5 – May 15). For “enhanced extraction” 280 µl of wastewater was extracted and eluted to 50 µl of elution buffer (May 19 – June 12). After June 12, 2020 viral RNA was extracted using chemagic Viral DNA/RNA 300 Kit special H96 (CMG-1033-S, Perkin Elmer) with the chemagic 360 (2024-0020, Perkin Elmer) automated platform. 300 µl of each sample was extracted according to manufacturer instructions and eluted in 100 µl sterile, nuclease-free water. All extracts were stored at −80 °C until quantification.

#### Rice nucleic acid extraction

Between June 22 and August 17, 2020, viral RNA was extracted using the AllPrep PowerViral DNA/RNA Kit (28000-50, Qiagen) with minimal modifications to manufacturer instructions. Immediately prior to bead beating, bead tubes containing the frozen filters were removed from the freezer and transferred to ice. 7 µL of □-Mercaptoethanol and 693 µL of PM1 solution were added to each bead tube. Sample tubes were bead beaten at max speed in a Mini-Beadbeater 24 (3,500 rpm; 112011, BioSpec) for 1 minute, returned to ice for 2 minutes, bead beaten for 1 minute, and then returned to ice. After bead beating, tubes were centrifuged at 17,000 g for 2 minutes to pellet the filter debris and beads. Approximately 450 µL of sample lysate from the bead beating tube was transferred into a rotor adapter for extraction using the QIAcube Connect (9002864, Qiagen) automated platform. Each sample was eluted in 50 µL nuclease-free water. All extracts were stored at −20 °C until quantification.

On August 24, the Rice laboratory switched to using the Maxwell 48 RSC automated platform (AS8500, Promega) for nucleic acid extractions. This decision to change extraction kits was based on a supply chain shortage of Qiagen AllPrep PowerViral kits. A head-to-head comparison of the Qiagen AllPrep PowerViral kit and Maxwell RSC PureFood GMO and Authentication Kit (AS1600, Promega) was performed by comparing N1 and N2 yields from 22 wastewater samples. We consistently observed significantly higher yields of SARS-CoV-2 RNA using the PureFood GMO kits (data not shown).

A modified protocol for the Maxwell RSC PureFood GMO kit, as recommended by Promega representatives, was used to extract samples on the Maxwell RSC 48. Bead tubes containing 0.1mm glass beads and sample filters were supplemented with 700 µL of CTAB. Sample tubes were bead beaten at max speed in a Mini-Beadbeater 24 (3,500 rpm; 112011, BioSpec) for 1 minute, returned to ice for 2 minutes, bead beaten for 1 minute, and then returned to ice. After bead beating, samples tubes were administered 40 µL of Proteinase K and then briefly vortexed to mix. Sample tubes were then incubated at 56 °C for 10 minutes followed by centrifugation for 2 minutes at 17,000 g. After centrifugation, 350 µL of supernatant was added to the first well of the Maxwell cartridge along with 300 µL of Lysis Buffer. The Maxwell RSC 48 completed the extraction and eluted the sample into 50 µL of elution buffer. Extracts were stored at −20 °C until quantification.

### Quantification of SARS-CoV-2 RNA

#### BCM quantification of SARS-CoV-2 RNA

The extracted RNA was tested using the CDC 2019-Novel coronavirus (2019-ncoV) Real-Time RT-PCR Diagnostic panel (*28*). The assay targets the nucleocapsid (N) gene (N1 and N2) of the SARS-CoV-2 genome. Real-time RT-PCR was performed using 10 µl of eluted RNA and 15 µl of TaqPath 1-step RT-PCR Master Mix, CG (A15299 Applied Biosystems) under the following cycling conditions: 25 °C for 2 minutes, 50 °C for 15 minutes, 95 °C for 2 minutes, and 45 cycles of 95 °C for 3 seconds, 55 °C for 30 seconds on a 7500 Fast Dx Real-Time PCR Instrument (4406985, Applied Biosystems) with SDS version 1.4 software. Samples were considered positive if N1, N2, Ct values were <40. The real-time RT-PCR included negative extraction, no template negative controls, and a standard curve of the linearized N plasmid to determine the genomic copy numbers of N1 and N2 in the samples. The standard curve ranged from 10,000-16 copies/mL with N1 primer values of R^2^: 0.992, efficiency: 99.1% and N2 primer values of R^2^: 0.969, efficiency: 97.4%. Limit of detection (LOD) was set as 2 gene copies/10µl RNA template. Applying a concentration factor of 30x from wastewater to RNA extract, and converting from µL to L, the LOD was calculated as 6,667 copies/L wastewater.

#### Rice quantification of SARS-CoV-2 RNA via RT-ddPCR

RNA extracts stored at −20 °C were thawed on ice, centrifuged at 17,000 g for 5 minutes to remove residual magnetic beads (if the Maxwell extraction platform was used, otherwise centrifugation is skipped), and supernatants were aliquoted into a 96 well plate. Reverse transcription and droplet digital PCR (ddPCR) were conducted with One-Step RT-ddPCR Advanced Kit for Probes (1864021, Bio-Rad) on the QX200 AutoDG Droplet Digital PCR System (1864100, Bio-Rad) to quantify the concentration of N1 and N2 SARS-CoV-2 gene targets in extracted samples. Limit of detection (LOD) was determined as 3 positive droplets per reaction well of ddPCR, which is equivalent to approximately 0.33 gene copies/µl RNA template. Applying a concentration factor of 1,000x from wastewater to RNA eluate (50mL wastewater is concentrated to generate a 50µl RNA extract), and converting from µl to L, the LOD was calculated as 330 copies/L wastewater.

The reaction composition followed the manufacturer recommendations. Each 22 µL duplex reaction contained 0.12 µL DNA-grade water, 5.5 µL Supermix, 2.2 uL reverse transcriptase, 1.1 µL DTT (15 mM), 0.55 µL N1 probe (0.25 µM), 0.5 µL N1 forward primer (0.9 µM), 0.5 µL N1 reverse primer (0.9 µM), 0.55 µL N2 probe (0.25 µM), 0.5 µL N2 forward primer (0.9 µM), 0.5 µL N2 reverse primer (0.9 µM), and 10 µL of RNA template (100 fg – 100 ng). The N1 and N2 sequences used here for primers and probes were taken from the recommended CDC sequences (**Table S2**). Primers and probes were purchased from Genewiz and Applied Biosystems. The RNA templates and the ddPCR plates were maintained on ice throughout the procedure. After droplet generation, thermocycling was conducted on a C1000 Touch Thermocycler (1851196, Bio-Rad). Thermocycling consisted of reverse transcription at 50 °C for 60 minutes, enzyme activation at 95 °C for 10 minutes, followed by 40 cycles of denaturation at 95 °C for 30 seconds and annealing/extension at 60 °C for 1 minute (2 °C/sec ramp rate), and completed with enzyme deactivation at 98 °C for 10 minutes. Samples were then held at 4 °C in the thermocycler for no more than 12 hours until droplet reading. Droplets were analyzed using automatic settings on the QuantaSoft v1.7.4 software, and manual thresholding was performed when automatic settings failed to detect clusters of positive and negative droplets.

#### Rice quantification of SARS-CoV-2 RNA via RT-Qpcr

One step reverse transcription and quantification was performed using TaqMan Fast Virus 1-Step Master Mix (4444434, Applied Biosystems). Each 10 µL duplex reaction contained 2.7 µL DNA-grade water, 2.5 µL master mix, 0.2 µL N1 probe (0.2 µM), 0.1 µL N1 forward primer (0.4 µM), 0.1 µL N1 reverse primer (0.4 µM), 0.2 µL N2 probe (0.2 µM), 0.1 µL N2 forward primer (0.4 µM), 0.1 µL N2 reverse primer (0.4 µM), and 4 0.2 µL N1 probe (0.2 µM), 0.1 µL N1 forward primer (0.4 µM), 0.1 µL N1 reverse primer (0.4 µM), and 4 µL RNA template assembled in Fast 96-well plates (Applied Biosystems). The N1 and N2 sequences used here for primers and probes were taken from the recommended CDC sequences (**Table S2**). Primers and probes were equivalent to those used in our RT-ddPCR. Thermocycling was completed in a QuantStudio 3 Real Time PCR System (A28567, Applied Biosystems) and consisted of reverse transcription at 50 °C for 5 minutes, enzyme activation at 95 °C for 20 seconds, and 40 cycles of denaturation at 95 °C for 3 seconds and annealing/extension at 60 °C for 30 seconds. Automatic background correction and thresholding were performed in QuantStudio Design and Analysis software (version 1.4). Linear DNA fragments of 250 bp each for N1 and N2 (Genewiz) were diluted in Herring Sperm DNA solution (D1811, Promega) and used as standards in a 10-fold dilution series of 5 concentrations from 28,000 copies/well to 2.8 copies/well in two replicates on every qPCR plate.

### RT-qPCR versus ddPCR quantification of SARS-CoV-2 RNA in influent wastewater samples

The Rice laboratory compared assays using RT-qPCR versus ddPCR for quantifying N1 and N2 targets in wastewater samples. To confirm that results generated between ddPCR and RT-qPCR corresponded, Rice directly compared concentrations of the same extracts as quantified by the two different methods. We found a relatively linear relationship (N1 - R^2^ = 0.80, N2 - R^2^ = 0.91), where RT-qPCR measurements were roughly 0.07 times and 0.31 times that of ddPCR for N1 and N2, respectively (**Fig. S1**). These results indicate that measurements from the two methods can be directly compared when an appropriate adjustment factor is applied. We also compared N1 to N2 concentrations as measured by the two methods. We found a linear relationship (ddPCR - R2 = 0.92, RT-qPCR = 0.93) where N2 measurements were about 0.7 times and 3.05 times N1 for ddPCR and RT-qPCR, respectively. Differences between ddPCR and qPCR, as well as N1 and N2 may be caused by differences in primer/probe binding efficiency between N1 and N2, biases introduced by standards in RT-qPCR, and/or differences in sensitivity of the different channels in ddPCR to detect their fluorophore of interest.

### Clinical positivity rate data

Per Governor’s Executive Order No. GA-10, all public and private entities conducting FDA-approved SARS-CoV-2 diagnostic tests are required to report all test results, including both positive and negative results, to the City of Houston Health Department (HHD). Currently, HHD utilizes the Houston Electronic Disease Surveillance System (HEDSS) to receive and store COVID-19 test results to perform daily surveillance and contact tracing activities for the City. There are multiple input methods of COVID-19 data into HEDSS from over 100 reporting entities across the U.S., including direct electronic laboratory records (ELRs), feed from several local hospitals/laboratories, batch uploads of file transfers received through SFTP sites, manual data entry of faxes, and ELRs from the Texas Department of State Health Services. A separate team audits data in HEDSS to determine if all test results for the City are reported to HHD in a timely manner. The lab audit team independently contacts and requests a separate data file of all COVID-19 results to compare with data currently in HEDSS from entities participating in the lab audit project. In addition, this team evaluates the completeness and quality of the required data fields received from the entity (e.g., percentage of results with race/ethnicity information or percentage of records with realistic specimen dates).

For WWTP activities, COVID-19 results are extracted from HEDSS and geocoded to wastewater treatment plant service area (WWTP) so that each test result has a corresponding WWTP (if within the city limits and can be geocoded). Daily counts of people tested for COVID-19 and people who have tested positive for COVID-19 are determined for each WWTP. This frequency data is used to calculate the positivity rate by WWTP service area.

### Data cleaning and early processing

Electronic data was updated weekly. It consisted of wastewater analysis results (N1 and N2 copies/L of wastewater) from BCM and Rice, and population and flow rate information from the City of Houston. In all cases, the information was at the WWTP level. The format of the files was comma separated files (CSV). The data was imported into R programming language through the RStudio Integrated Development Environment (IDE). Several steps were performed to make sure that naming conventions as well as sample identification were consistent between the diverse data providers. The processed weekly dataset was appended to the historical dataset. For analysis purposes, this dataset was further enriched with the historical daily report of total of persons tested and number of positive cases by WWTP, as well as the daily and 14-day moving average positivity rate. Wastewater loads were calculated by multiplying the viral concentration data by the average influent flow rate in L/day. Measurements falling below the LOD provided by the respective lab where imputed based on a random selection between ½ of the LOD and the LOD. Wastewater viral load data was log transformed so results could be expressed as log 10 copies/day. Finally, data was aggregated by date and WWTP, to be used as input for the statistical analysis.

### Statistical methods and model

A multilevel regression model was fit to the wastewater viral load data in log10 copies/day obtained from each lab. The base model included fixed effects for WWTP, lab, targets of N1 and N2, and a random effect to incorporate the three replicates of each combination. To assess a one-week trend, a regression model using the current and previous week was used to calculate the week-to-week change for each WWTP (see **Fig 4B**). A multilevel model was used, which included and indicator variable representing week n-1 and week n interacted with the WWTP to assess the change, lab, targets of N1 and N2, and a random effect to incorporate the three replicates of each combination.

In addition to the weekly trend assessment, nonparametric regression methods were implemented to obtain the longitudinal profile for the measured viral load and clinical positivity rate for each WWTP and the city as a whole. A semi-parametric spline regression model was used to smooth the weekly aligned viral load in log10 copies/day from each lab for each WWTP. The individual estimates for each WWTP were aggregated to obtain the longitudinal trend for the region. For the total viral load, the log10 transformation was reversed for the smoothed trends of each WWTP, the estimates were summed and then the log10 transformation was again applied. For the average trend across WWTP, the smoothed estimates were averaged. Variance estimates are obtained by summing the individual variances across WWTP, with an implicit assumption of little to no correlation across WWTP. Similarly, a semi-parametric spline regression model was used to smooth the clinical positivity rate for each WWTP, and the overall clinical positivity rate for the region.

Parametric and semi-parametric regression models were used to adjust for changes in methodologies within and between labs. The first transition involved the Baylor lab changing viral concentration methods from PEG precipitation to electronegative filtration (HA). Based on the head-to-head measurements of 15 wastewater samples, PEG measurements were adjusted to HA measurements. A simple linear regression model was fit to each of the N1 and N2 targets of the log10 copies/L from HA, with the corresponding log10 PEG copies/L as the predictor variable. The N1 regression yielded an intercept of 3.24 (p=0.04), slope of 0.81(p<0.0001) and adjusted R-squared of 0.678. The N2 regression yielded an intercept of 4.65 (p=0.07), slope of 0.67 (p=0.01) and adjusted R-squared of 0.3419. Predicted values from these regression models formed the adjusted N1 and N2 copies from May 11 through June 29.

An alignment of measurements between Rice and Baylor was performed weekly, prior to obtaining the nonparametric longitudinal estimates. A semi-parametric regression model fitting Baylor N1 and N2 average log10 copies/L with Rice N1 and N2 average log10 copies/L is obtained. The model includes an indicator variable for a Rice nucleic acid extraction method change on August 24 times a natural spline with 8 degrees of freedom. The fitted model had an adjusted R-squared of 0.61 and improved each week as additional observations were used in the adjustment. Rice average N1 and N2 log10 copies/L were adjusted to Baylor levels using the fitted regression model.

Since the methodology for longitudinal trends is based on splines, missing values may inappropriately impact the estimate. An additional data preparation step, was to linearly interpolate up to 2 missing values in the log10 copies/day temporal series for each WWTP, as well as the positivity rate series when possible.

Spearman rank based correlation was obtained between the smoothed WWTP viral load estimates and the clinical positivity rate based on the estimates for Monday of each week of our study period. In addition, the Spearman correlation was obtained for the raw (unsmoothed) series. A cross-correlation analysis was performed using the smoothed wastewater viral load and smoothed positivity rate series. Spearman cross-correlations were computed with wastewater leading positivity rate between 0 and 14 days. On average, the cross-correlations were around 0.8 over the 14-day period, demonstrating that wastewater viral load estimates represent a leading indicator of clinical positivity rate.

Predictive models were explored for the clinical positivity rate based on the wastewater viral load for each WWTP service area. The predictive models used were regression models with different predictor variables derived from the longitudinal estimates of the wastewater viral load (in log10 copies day^-1^) for each WWTP. The models were as follows:

*Model 1:* Indicator for WWTP, current, 1-week and 2-week lag of log10 copies/day,

*Model 2:* Indicator for WWTP, 1-week and 2-week lag of log10 copies/day

Each model incorporated the time series correlation structure through an autoregressive model of order 1 for the error process. The autoregressive parameter estimates were 0.516 and 0.490 for Models 1 and 2, respectively.

The models were fit using generalized least squares to incorporate the autoregressive error structure, using R-function gls in the nlme package (*29*). In-sample comparisons demonstrated strong predictive ability; the Spearman correlation between predicted PR and smoothed PR is 0.83 for model 1 and 0.79 for model 2. As expected, Model 1 is a stronger model since it used current wastewater values. To quantitatively access the strength, we computed the Bayesian Information Criterion (BIC) for nested models. The model with the lowest BIC provides stronger characterization between wastewater viral loads and PR. The BIC for Model 1 = −75.08, while BIC Model 2 = 15.67, illustrating the greater explanatory power of Model 1. Summary model results are presented in **Table S3**.

In Models 1 and 2, the intercept represents the contribution from the WWTP 69^th^ Street, the WWTP serving the largest percentage of the Houston population. For both fitted models, the coefficients of the other 34 WWTP are significantly different from 0 (p<0.001) with the exception of Almeda Sims, Southwest, and Upper Brays Bayou, indicating these three WWTPs behaved similarly to the 69^th^ Street WWTP.

Predicted positivity rates were obtained for each WWTP by evaluating Model 1 and Model 2 at the smoothed estimates of wastewater viral load in log10 copies/day. Time series plots for all WWTPs, are provided in **Figure S4**.

## Data Availability

Data has been submitted for publication on the Kinder Institute Urban Data Platform (https://www.kinderudp.org/). Once accepted, data will be issued a DOI and data will be accessible following government procedures.

## Acknowledgments

We thank Paul Zappi, Rae Mills, Carol LaBreche, Walid Samarneh, and Aisha Niang from Houston Water for their assistance in collecting wastewater samples. We thank Braulio Garcia, Courtney Hundley, Jeremy Rangel, Kelsey Caton, Rebeca Schneider, Daniel Bahrt, Kaavya Damakonda, Patrick Key, and Naomi Macias from the Houston Health Department for their assistance in sample collection and data analysis. We thank Jessica Chen, Kristina Cibor, Esther Lou, Basmah Maiga, Camille McCall, Dylan Nguyen, Pavan Raja, Kiara Reyes Gamas, Naomi Senehi, and Emma Zohner and for their assistance in sample collection, processing, analysis, and project management. We thank Jeseth Delgado Vela, Adam Smith, Nadine Kotlarz, Francis de los Reyes, and Angela Harris for their discussions on wastewater-based epidemiology.

## Funding

This work was supported by the Houston Health Department and grants from the National Science Foundation (CBET 2029025) and seed funds from Rice University and Baylor College of Medicine. P.K. was funded by a Johnson & Johnson WiSTEM2D award. Z.W.L. was funded by an Environmental Research & Education Foundation scholarship and Rice University. P.A. was funded by a National Science Foundation award (CBET 1932000). T.C. was funded by the National Academies of Science, Engineering, and Medicine Gulf Research Early Career Research Fellowship.

## Notes

### Competing Interest Statement

The authors have declared no competing interest.

