## Supplementary Materials for "Wastewater Analysis of SARS-CoV-2 as a Predictive Metric of Positivity Rate for a Major Metropolis"

This file includes:

Figs. S1 to S5

Tables S1 to S3

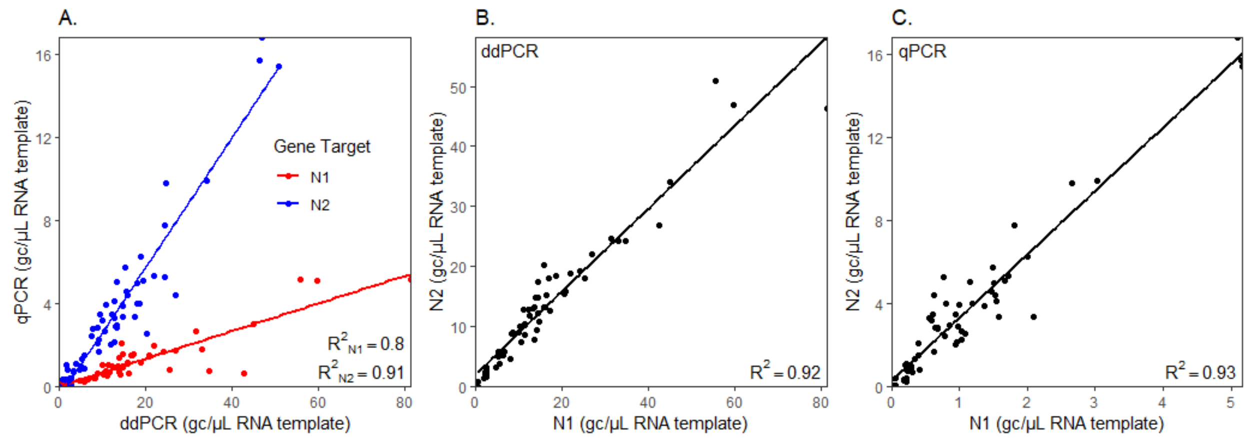


**Fig. S1.** (**A**) Comparison between RT-ddPCR and RT-qPCR of either N1 measurements or N2 measurements in gene copies (GC) per µL of RNA extract template. (**B**) Comparison of N1 to N2 as measured by RT-ddPCR. (**C**) Comparison of N1 to N2 as measured by RT-qPCR.

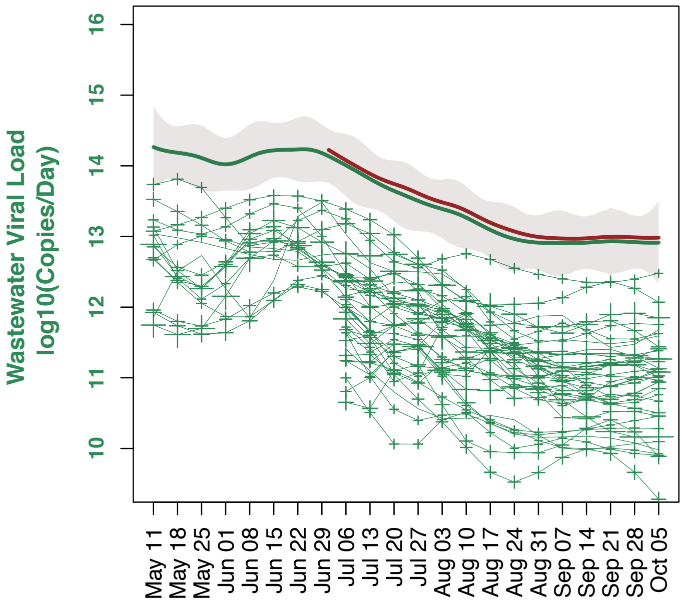


**Fig. S2.** Each + indicates a sample collected from a WWTP and those connected with lines were collected from the same WWTP on different weeks. The size of the + denotes the level of uncertainty for each set of observations. A nonlinear regression (spline) was fit to the observations from each WWTP. Sixteen WWTPs were sampled since May 11, and the remaining 23 WWTPs were sampled starting on July 6. The individual splines of the original 16 WWTPs were inverse log10 transformed, summed, and then 1og10 transformed to form the overall spline (green line). The grey represents the 95% confidence band for the overall estimate and is derived from the sum of the variances of each spline. The red line is the aggregate spline of the viral loads from all 39 WWTPs.

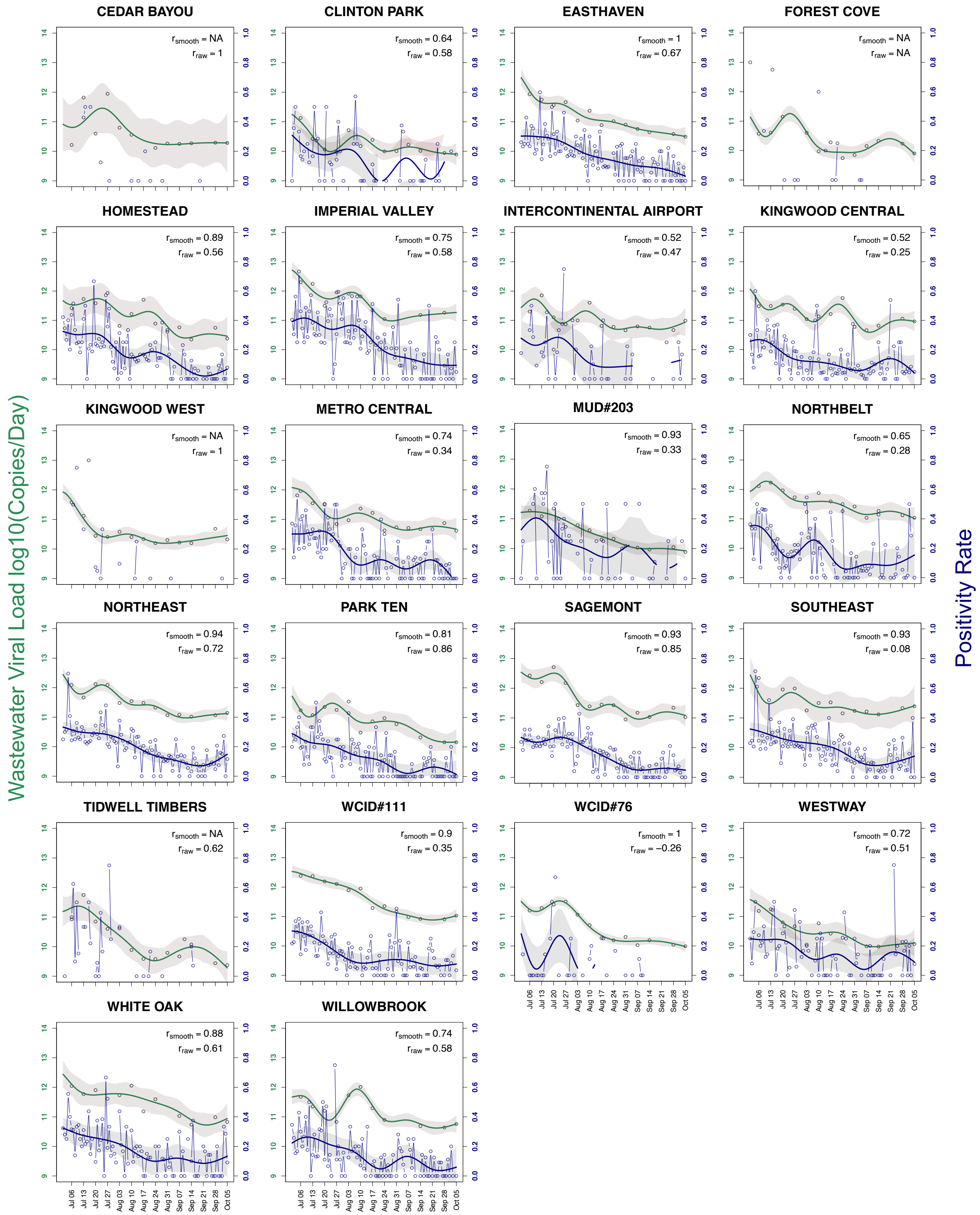


**Fig. S3.** Wastewater viral loads and positivity rates for individual WWTPs sampled between July 6 and October 5. Individual observations for the wastewater viral load and positivity rate are denoted by ◦ and green (wastewater) and blue (positivity rate) lines are the nonlinear regressions (splines) fit to the observations. Grey represents the 95% confidence bands. $r_{smooth}$ is the r Spearman estimate between the wastewater and positivity rate splines taken weekly, and $r_{raw}$ is between the raw observations taken weekly on the dates with wastewater observations. Positivity rates were only calculated if there were at least 4 clinical tests performed in that sewershed population on that day.

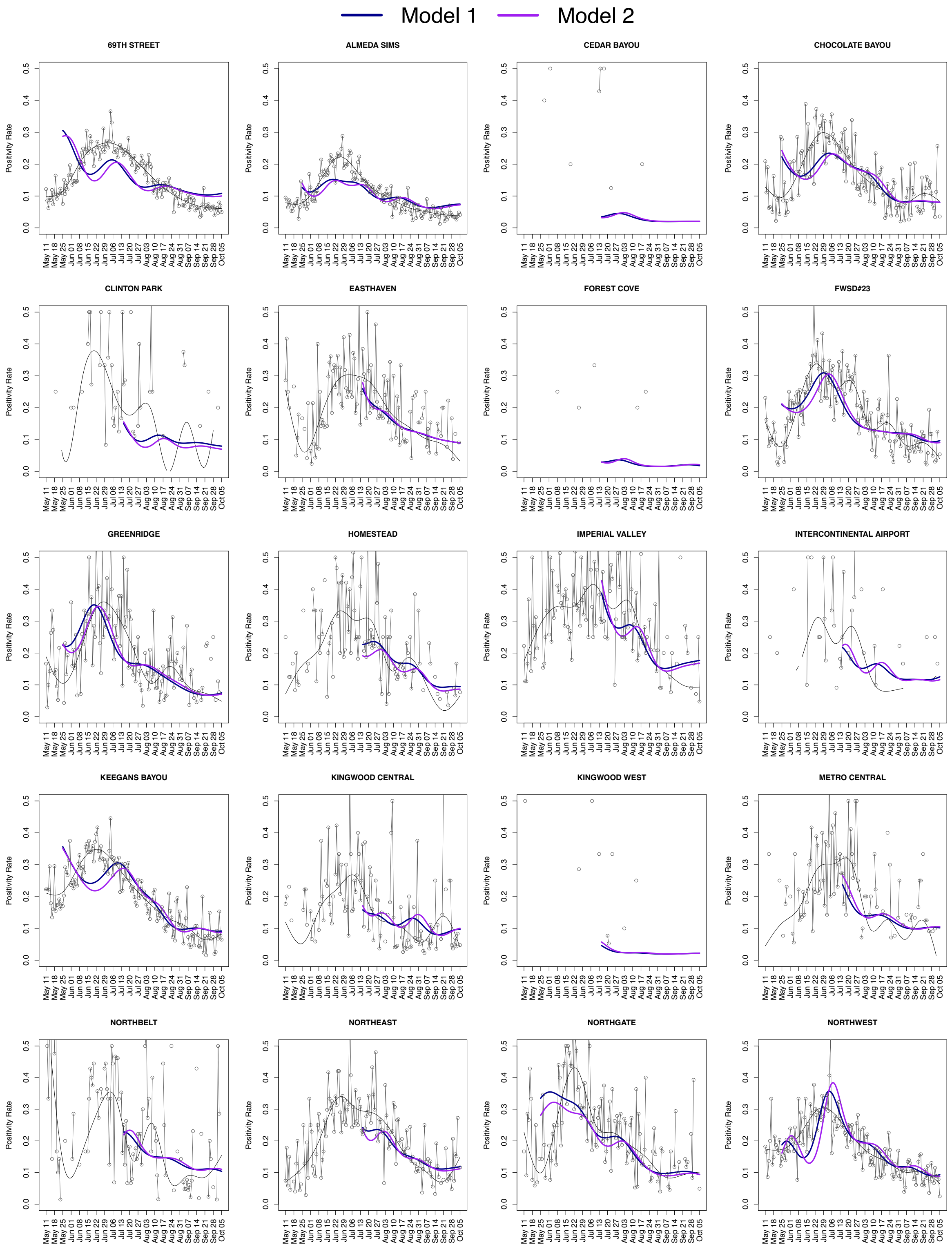
Model 1
Model 2
69TH STREET
ALMEDA SIMS
CEDAR BAYOU
CHOCOLATE BAYOU
CLINTON PARK
EASTHAVEN
FOREST COVE
FWSD#23
GREENRIDGE
HOMESTEAD
IMPERIAL VALLEY
INTERCONTINENTAL AIRPORT
KEEGANS BAYOU
KINGWOOD CENTRAL
KINGWOOD WEST
METRO CENTRAL
NORTHBELT
NORTHEAST
NORTHGATE
NORTHWEST
Positivity Rate
May 11 May 18 May 25 Jun 01 Jun 08 Jun 15 Jun 22 Jun 29 Jul 06 Jul 13 Jul 20 Jul 27 Aug 03 Aug 10 Aug 17 Aug 24 Aug 31 Sep 07 Sep 14 Sep 21 Sep 28 Oct 05

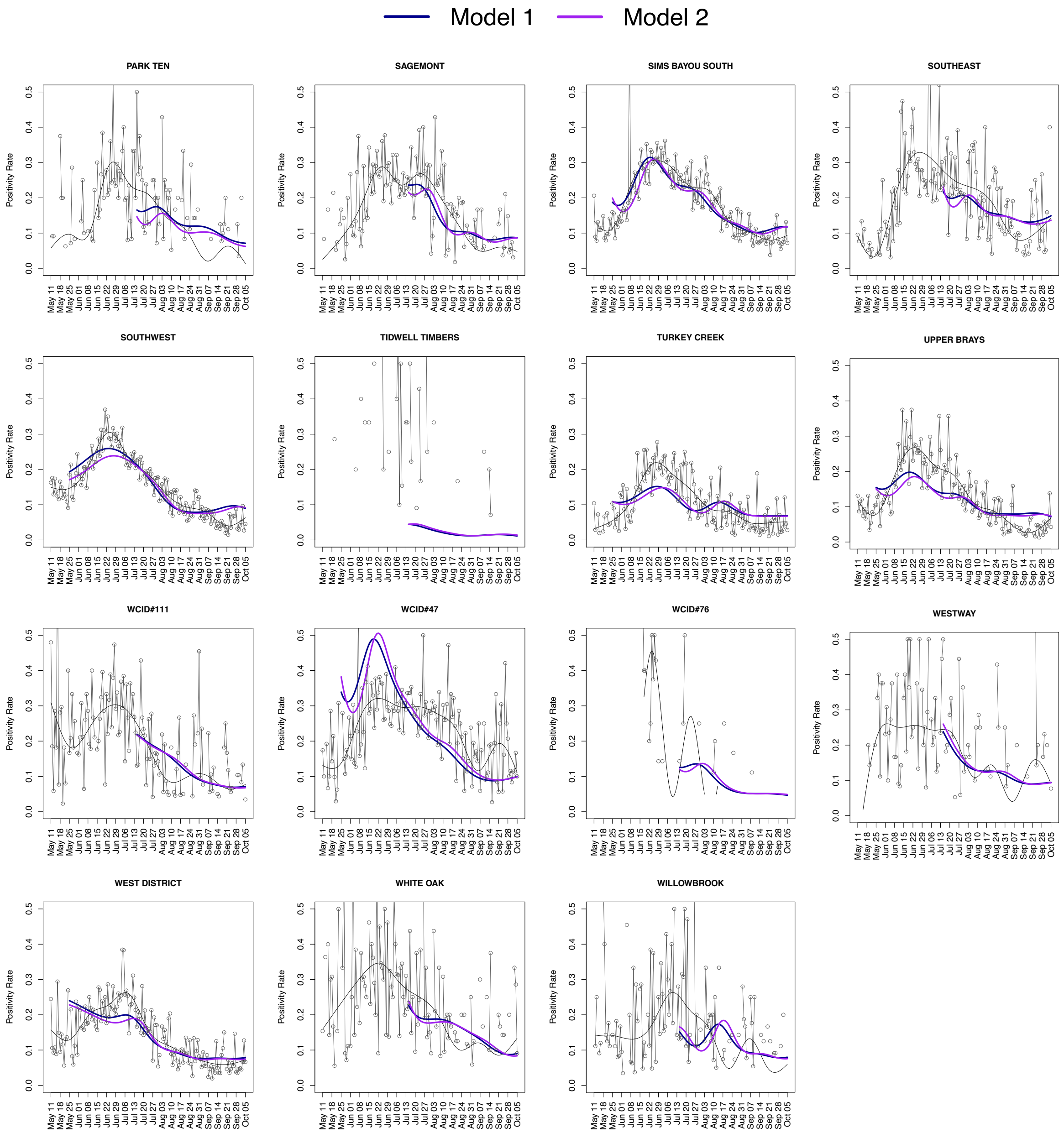


**Fig. S4.** Comparison of predicted positivity rates using model 1 (blue) and 2 (purple) against clinical positivity rates for the sewershed. Daily clinical positivity rates are shown as grey circles and smoothed positivity rate is represented by the grey lines. Positivity rates were only calculated if there were at least 4 nasal clinical tests performed in that sewershed population on that day.

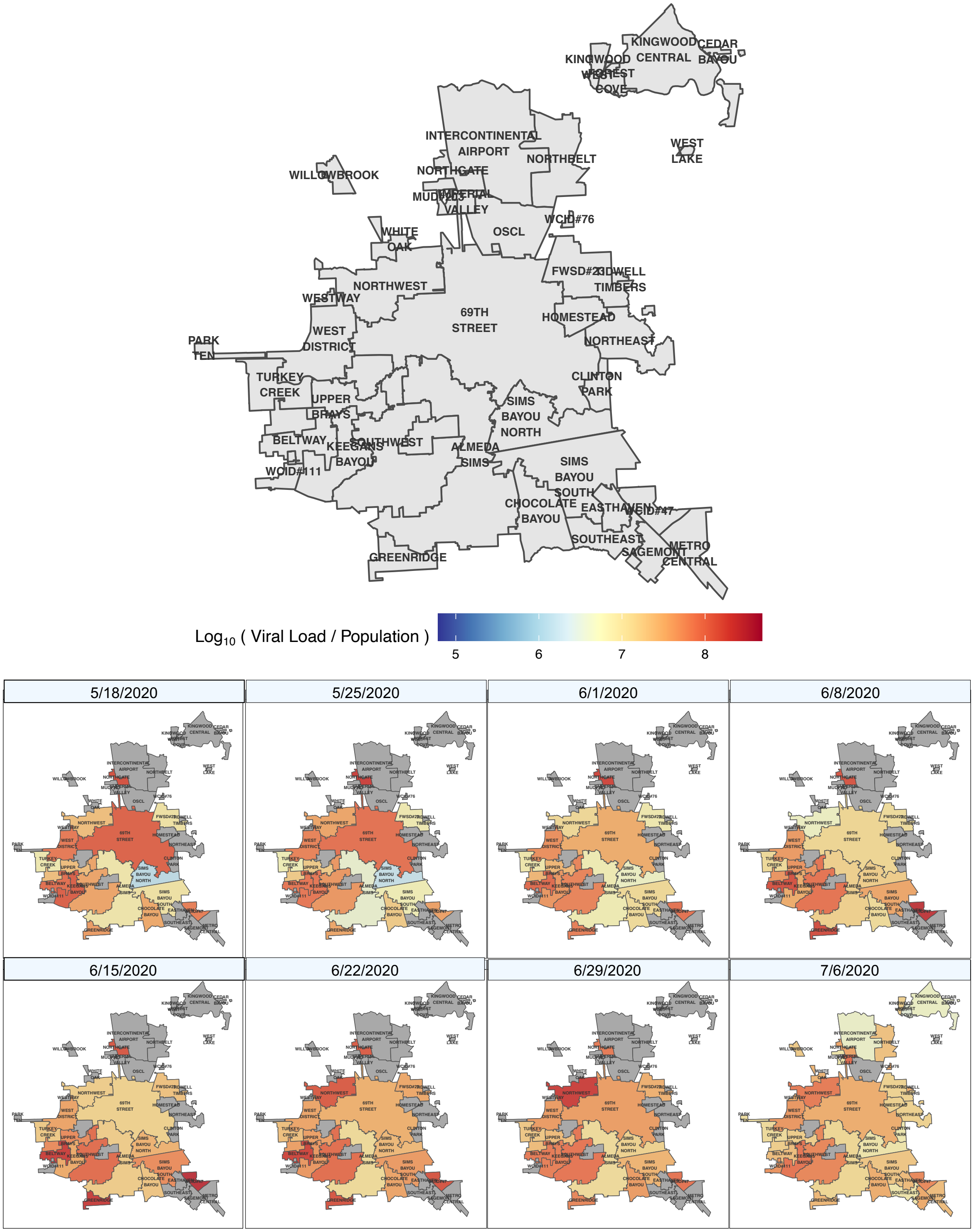
KINGWOOD
CEDAR BAYOU
KINGWOOD
CENTRAL
WEST FOREST
COVE
INTERCONTINENTAL
AIRPORT
NORTHBELT
WEST LAKE
WILLOWBROOK
NORTHGATE
MUDDY
IMPERIAL VALLEY
WCID#76
WHITE OAK
OSCL
FWSD#23
IDLEWELL TIMBERS
NORTHWEST
WESTWAY
69TH STREET
HOMESTEAD
WEST DISTRICT
PARK TEN
NORTHEAST
TURKEY CREEK
CLINTON PARK
UPPER BRAYS
SIMS BAYOU NORTH
BELTWAY
KEEGANS BAYOU
SOUTHWEST
ALMEDA SIMS
SIMS BAYOU SOUTH
WCID#111
CHOCOLATE BAYOU
EASTHAVEN
WCID#47
SOUTHEAST
METRO CENTRAL
SAGEMONT
GREENRIDGE
Log10 ( Viral Load / Population )
5
6
7
8
5/18/2020
5/25/2020
6/1/2020
6/8/2020
6/15/2020
6/22/2020
6/29/2020
7/6/2020

56

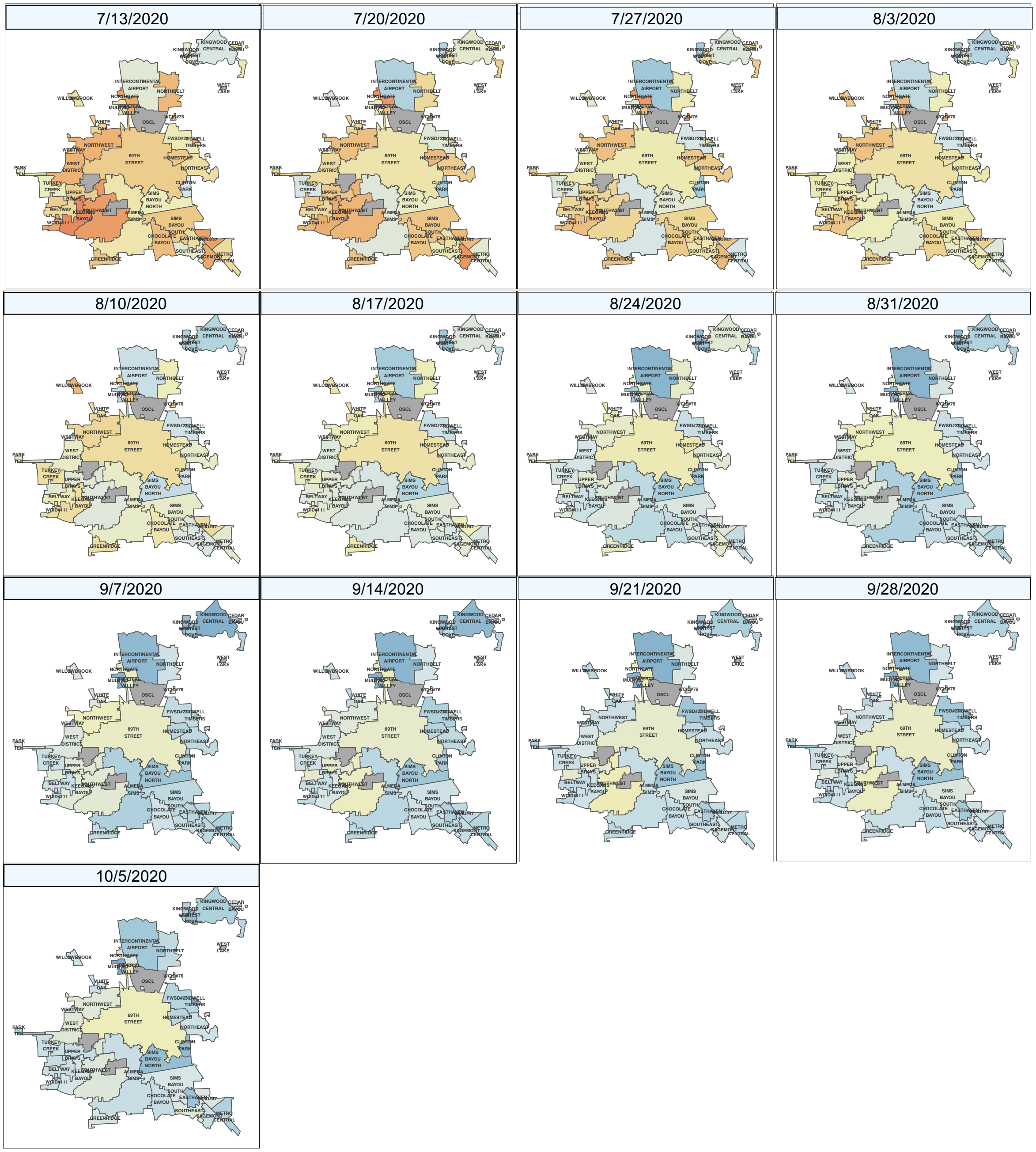


**Fig. S5.** Maps of population-normalized viral load for each sewershed (outlined in black) for each week between May 11 and October 5 based on regression model estimates of viral load and normalized using the service area populations in Table S1.

**Table S1.** Wastewater treatment plants sampled, average flow rates, service populations, and geographic service areas

| Wastewater treatment plant | Abbreviation | Flowrate, MGD (AVG ± SD) | Population | Average gal/cap/day | Area, square miles |
|---|---|---|---|---|---|
| 69th Street | 69 | 80.03 ± 21.77 | 563,727 | 129 | 96.72 |
| Almeda Sims | AS | 13.76 ± 11.93 | 330,126 | 34 | 54.93 |
| Beltway | BW | 6.93 ± 4.70 | 134,557 | 47 | 9.76 |
| Cedar Bayou | CD | 0.78 ± 0.42 | 15,601 | 44 | 3.27 |
| Chocolate Bayou | CB | 4.03 ± 4.19 | 64,306 | 54 | 14.61 |
| Clinton Park | CP | 0.69 ± 0.81 | 14,295 | 40 | 4.14 |
| Easthaven | EH | 1.89 ± 1.85 | 46,111 | 33 | 4.78 |
| FWSD#23 | 23 | 3.09 ± 2.52 | 60,791 | 43 | 15.14 |
| Forest Cove | FC | 0.28 ± 0.13 | 13,447 | 19 | 2.73 |
| Greenridge | GR | 3.06 ± 3.06 | 54,739 | 46 | 6.87 |
| Homestead | HO | 1.58 ± 1.53 | 23,087 | 55 | 6.12 |
| Imperial Valley | IV | 1.75 ± 0.70 | 28,771 | 56 | 2.22 |
| Intercontinental Airport | IA | 1.91 ± 0.63 | 110,793 | 16 | 38.73 |
| Keegans Bayou | KB | 14.25 ± 10.31 | 184,339 | 66 | 13.78 |
| Kingwood Central | KW | 3.49 ± 1.46 | 94,774 | 36 | 23.04 |
| Kingwood West | MG | 0.61 ± 0.20 | 28,972 | 19 | 2.6 |
| MUD#203 | 203 | 0.38 ± 0.12 | 22,558 | 16 | 2.57 |
| Metro Central | MC | 1.99 ± 1.64 | 44,793 | 36 | 9.86 |
| Northbelt | NO | 2.37 ± 1.49 | 70,168 | 32 | 15.79 |
| Northeast | NE | 3.88 ± 4.25 | 68,316 | 190 | 14.41 |
| Northgate | NG | 2.75 ± 1.03 | 48,292 | 52 | 3.6 |
| Northwest | NW | 9.99 ± 5.84 | 147,177 | 61 | 22.62 |
| Park Ten | PT | 0.62 ± 0.31 | 17,889 | 30 | 2.19 |
| Sagemont | SG | 4.52 ± 3.49 | 67,929 | 56 | 5.9 |
| Sims Bayou South* | SS | 23.93 ± 18.22 | 304,253 | 68 | 47.84 |
| Sims Bayou North* | SB | 8.22 ± 7.54 | 304,253 | 22 | 47.84 |
| Southeast | SE | 4.88 ± 4.85 | 86,598 | 45 | 9.06 |
| Southwest | SW | 37.59 ± 26.39 | 395,901 | 80 | 38.72 |
| Tidwell Timbers | TT | 0.11 ± 0.06 | 6,564 | 15 | 0.57 |
| Turkey Creek | TC | 7.00 ± 4.85 | 91,460 | 65 | 10.46 |
| Upper Brays | UB | 10.33 ± 7.44 | 167,098 | 52 | 12.81 |
| WCID#111 | 111 | 2.24 ± 0.28 | 57,865 | 38 | 3.35 |
| WCID#47 | 47 | 3.36 ± 2.28 | 46,775 | 63 | 6.27 |
| WCID#76 | 76 | 0.37 ± 0.22 | 5,130 | 62 | 0.5 |
| West District | WD | 10.06 ± 6.62 | 126,321 | 67 | 17.86 |
| West Lake | WL | 0.20 ± 0.07 | 35,769 | 5 | 0.53 |
| Westway | WW | 0.40 ± 0.18 | 8,121 | 48 | 0.99 |
| White Oak | WO | 1.84 ± 0.91 | 48,203 | 35 | 3.31 |
| Willowbrook | WB | 1.28 ± 0.52 | 31,072 | 36 | 3.01 |
| **TOTAL** | | **272.23 ± 159.04** | **3,666,688** | **49±32** | **532** |

*Sims Bayou South and North have overlapping geographic service areas.

**Table S2.** CDC primers for SARS-CoV-2 N1 and N2

| **Name** | **Description** | **Sequence** (5' - 3') |
|---|---|---|
| CoV2_N1-F | CoV2 N1 Forward Primer | GACCCCAAAATCAGCGAAAT |
| CoV2_N1-R | CoV2 N1 Reverse Primer | TCTGGTTACTGCCAGTTGAATCTG |
| CoV2_N1-Pr | CoV2 N1 Probe | VIC – ACCCCGCATTACGTTTGGTGGACC – QSY |
| CoV2_N2-F | CoV2 N2 Forward Primer | TTACAAACATTGGCCGCAAA |
| CoV2_N2-R | CoV2 N2 Reverse Primer | GCGCGACATTCCGAAGAA |
| CoV2_N2-Pr | CoV2 N2 Probe | FAM – ACAATTTGCCCCCAGCGCTTCAG – QSY |

**Table S3**. Summary model results for predictive Model 1 and 2.

| Model 1 | DF | F-Value | p-value |
|---|---|---|---|
| Intercept | 1 | 12183.16 | < 0.0001 |
| WWTP | 33 | 5.75 | <0.0001 |
| Current log10 copies/day | 1 | 431.82 | <0.0001 |
| 7-day lead of viral load in log10 copies/day | 1 | 70.85 | <0.0001 |
| 14-day lead of viral load in log10 copies/day | 1 | 26.64 | <0.0001 |

| Model 2: | DF | F-Value | p-value |
|---|---|---|---|
| Intercept | 1 | 10672.51 | < 0.0001 |
| WWTP | 33 | 4.91 | <0.0001 |
| 7-day lead of viral load in log10 copies/day | 1 | 422.14 | <0.0001 |
| 14-day lead of viral load in log10 copies/day | 1 | 46.26 | <0.0001 |
